# Alzheimer’s disease and the therapeutic potential of theta burst stimulation: A systematic review of preclinical and clinical studies

**DOI:** 10.1101/2025.09.15.25335839

**Authors:** Negin Eissazade, Pouria Akbarikoli, Neguine Rezaii, Mohammadreza Shalbafan, Hesam Mosavari

**Author notes:** Corresponding author: ^1^Negin Eissazade, Instutional address: Cellular and Molecular Research Center, Iran University of Medical Sciences, Hemmat Highway, Tehran, Iran., ^2^Hesam Mosavari, Institutional address: Artificial Intelligence in Health Research Center, Iran University of Medical Sciences, Hemmat Highway, Tehran, Iran.

## Abstract

**Introduction:** Alzheimer’s disease (AD) is a progressive neurodegenerative disorder characterized by cognitive decline and behavioral impairment. Despite growing research efforts, effective disease-modifying interventions remain elusive. Theta burst stimulation (TBS), a form of repetitive transcranial magnetic stimulation (rTMS), has shown promise in modulating cortical excitability, synaptic plasticity, and neuroprotective mechanisms. This systematic review aimed to evaluate the efficacy and safety of TBS in AD by synthesizing findings from preclinical and clinical studies.

**Methods:** A systematic search of PubMed, Scopus, Embase, Web of Science, and the Cochrane Library was conducted up to January 2025. Studies investigating TBS, including intermittent TBS [iTBS] and continuous TBS [cTBS], in human patients with AD or in animal models of AD were included. Primary outcomes included cognitive function and neuropsychiatric symptoms. Secondary outcomes included biomarkers of neuroplasticity and neurodegeneration. The risk of bias was assessed using the Joanna Briggs Institute (JBI) and Systematic Review Centre for Laboratory Animal Experimentation (SYRCLE) tools.

**Results:** Twenty studies met the inclusion criteria, comprising six preclinical and 14 clinical studies.

Preclinical evidence suggests that TBS reduces amyloid-beta deposition, enhances synaptic plasticity, mitigates neuroinflammation and oxidative stress, and improves cognitive performance in animal models of AD. iTBS targeting the dorsolateral prefrontal cortex (DLPFC) improved cognitive function, depression, anxiety, and activities of daily living, with some studies reporting non-significant findings in specific neuropsychological assessments. Neuroplasticity changes were observed in motor-evoked potentials and resting motor thresholds, with lower responses in patients with AD and variable reproducibility over time. Additionally, structural and functional brain changes, including preserved hippocampal volume and enhanced frontal beta activity, were associated with cognitive improvements. Adverse events were mild and well-tolerated.

**Conclusion:** Preclinical studies robustly support the neuroprotective and neuropsychiatric effects of TBS in AD models. Clinical findings suggest possible benefits of TBS, particularly iTBS over the DLPFC; however, clinical results are inconsistent and limited by small sample sizes and protocol variability. These findings highlight the challenges of translating animal data to humans and underscore the need for larger, controlled randomized clinical trials to confirm safety and efficacy and optimize treatment parameters.

## 1. Background

Alzheimer’s disease (AD) is the leading cause of dementia worldwide, with its prevalence projected to escalate significantly in the coming decades (1). This progressive neurodegenerative disorder is characterized by cognitive and behavioral deterioration resulting from widespread degenerative changes within the central nervous system (1). Despite substantial advances in pharmacological research, effective therapies capable of halting or reversing AD progression remain elusive. While new generations of FDA-approved medications have been shown to slow disease progression, they are associated with potentially serious side effects, necessitating careful risk-benefit assessments (2, 3). Consequently, alternative and complementary treatment modalities are garnering increasing attention (1, 4).

Theta burst stimulation (TBS), a form of repetitive transcranial magnetic stimulation (rTMS), was first introduced in 2005 and has since emerged as a promising intervention for modulating cortical excitability and synaptic plasticity (5, 6). Unlike traditional rTMS protocols that utilize fixed high-or low-frequency magnetic pulses, TBS delivers stimulation in short bursts, typically around 190 seconds, resulting in prolonged cortical excitability enhancement (7). TBS mimics the natural activity of the brain during learning, employing bursts of three biphasic pulses at 50 Hz, separated by 5 Hz (200-millisecond intervals) (6). This innovative design allows for shorter treatment sessions, reduced stimulation intensities, and comparable safety and efficacy to conventional rTMS (5). TBS can be administered in two primary modes: continuous TBS (cTBS) and intermittent TBS (iTBS). cTBS, delivered over 20–40 seconds, induces long-term depression and reduces cortical excitability, suggesting a possible dependence on NMDA (N-methyl-D-aspartate) receptor activity. In contrast, iTBS is administered in two-second trains interspersed with eight-second intervals, over a total duration of 190 seconds, facilitating long-term potentiation (LTP) and enhancing cortical excitability (5, 6). Importantly, variations in protocol parameters, such as pulse number, and stimulation site (e.g., dorsolateral prefrontal cortex [DLPFC], primary motor cortex [M1], and cerebellum) can modulate or even reverse the expected effects, highlighting the adaptability of TBS protocols (5, 6, 8, 9). The clinical applications of TBS, particularly iTBS, have demonstrated transformative potential. While initially validated for treatment-resistant depression (10), iTBS is increasingly recognized for its benefits in neurodegenerative conditions, including AD, Parkinson’s disease, and multiple sclerosis (11, 12). Furthermore, accelerated iTBS (aiTBS) delivers the same total pulse count across multiple daily sessions, this not only induces long-lasting cortical excitability, but also reduces clinic visits and offers patients quicker relief while maintaining comparable symptom improvement (13–16).

Given its non-invasive nature and capacity to target specific neural substrates, TBS holds significant promise as a novel therapeutic approach in AD. This systematic review aims to comprehensively evaluate the efficacy and safety of TBS in AD by synthesizing evidence from the existing clinical trials and experimental studies.

## 2. Materials and methods

The study protocol was registered on the International Prospective Register of Systematic Reviews (PROSPERO) [ID: CRD42025632613]. The study is reported in accordance with the guidelines outlined in the Preferred Reporting Items for Systematic Reviews and Meta-Analyses (PRISMA) (17).

### 2.1. Eligibility criteria

We used the PICOT framework (Population, Intervention, Comparator, Outcomes, and Timeframe) to structure this review and specifically address the research question: *“Is theta burst stimulation safe and effective for improving the symptoms associated with AD?”.* Eligible studies included preclinical animal models of AD, such as APP/PS1 transgenic mice and Wistar rats subjected to streptozotocin (STZ) or trimethyltin (TMT)-induced neurodegeneration, as well as human adults diagnosed with probable AD based on established clinical criteria (18–22), some supported by biomarker confirmation. The intervention comprised TBS protocols applied either as standalone treatments or as components of rTMS for therapeutic purposes or physiological assessment. Comparators included wild-type mice matched to APP/PS1 transgenic models, Wistar rats receiving saline injections instead of neurotoxins, placebo or sham stimulation groups exposed to noise or manual handling without active TBS, and, in clinical studies, sham stimulation, standard care, other neuromodulatory interventions, untreated controls, or healthy controls. Outcomes assessed encompassed a broad range of clinical and neurobiological measures evaluated through validated and standardized scales. In preclinical studies, primary outcomes encompassed cognitive and behavioral assessments (e.g., Morris water maze, Y-maze), along with a comprehensive set of cellular and molecular markers, including Aβ deposition, astrocyte and microglial activation (GFAP, Iba1, CD68), neuronal integrity and survival (NeuN, MAP-2), synaptic plasticity (PSD95, SNAP25, SYN1), apoptotic signaling (Caspase-3, Bax/Bcl-2 ratio, TUNEL), oxidative stress markers (SOD, MDA, GSH), neurotrophic support (BDNF), and glymphatic function (aquaporin-4). In clinical studies, the primary outcomes focused on changes in clinical symptoms measured by standardized clinical scales, including: Activities of Daily Living (ADL); Alzheimer’s Disease Assessment Scale-Cognitive Subscale (ADAS-Cog); ALBA Screening Instrument (ASI); Auditory Verbal Learning Test (AVLT); Boston Naming Test (BNT); Clock-Drawing Test (CDT); Clinical Dementia Rating (CDR); Color Trail Test (CTT); Digit Span Test (DST); Hamilton Anxiety Rating Scale (HAMA); Hamilton Depression Rating Scale (HAM-D or HDRS); Hooper Visual Organization Test (HVOT); Judgment of Line Orientation (JLO) test; Logical Memory Test (LMT); Mini-Mental State Examination (MMSE); Montreal Cognitive Assessment (MoCA); Neuropsychiatric Inventory (NPI); Repeatable Battery for the Assessment of Neuropsychological Status (RBANS); Self-Rating Depression Scale (SDS); Stroop Color and Word Test (SCWT); Symbol Digit Modalities Test (SDMT); Trail Making Test (TMT); and Verbal Fluency Test – Semantic and Letter Fluency (VFT-S/L). Other outcomes related to underlying AD pathologies included changes in biomarkers (e.g., *BDNF*), brain connectivity measures, and electrophysiological assessments of neuronal excitability. The timeframe of assessment varied across studies, covering both acute and longitudinal effects of TBS. Only studies published in English were included.

Studies were excluded if they investigated non-Alzheimer’s dementias or unrelated neurological conditions, or lacked original experimental data. Non-primary research publications, including reviews, editorials, commentaries, book chapters, study protocols, and non–peer-reviewed materials such as conference abstracts without full data, were excluded. Additionally, studies were excluded if they failed to report relevant clinical, cognitive, behavioral, neurobiological, or biomarker outcomes measured by validated and standardized instruments. Studies with inadequate methodological rigor or assessed as high risk of bias were also excluded.

### 2.2. Information sources, search strategy, and study selection

Potentially eligible articles were identified through an electronic search of bibliographic databases (PubMed, Scopus, Embase, Web of Science, and Cochrane Library) in January 2025. No restrictions were applied regarding publication date. The search strategy included the terms “*Theta Burst Stimulation*,” and “*Alzheimer’s Disease*,” along with related keywords, which were combined using Boolean operators in the free-text search fields. The detailed search strategy is provided in Supplementary Material 1.

Studies were initially screened based on titles and abstracts. Articles that were clearly irrelevant or did not meet the inclusion criteria were excluded. In cases where eligibility was unclear, a full-text review was performed. Additionally, the reference lists of included articles were manually reviewed to identify any potentially relevant studies not captured through the database search; however, this process did not yield any additional articles.

Screening, study selection, data extraction, and risk of bias assessment were conducted independently by two reviewers and cross-checked for accuracy. Any discrepancies between reviewers were resolved through discussion, or consultation with a third reviewer.

### 2.3. Data collection and synthesis

Extracted data included study design, sample size, subject characteristics (e.g., species, strain, age, and sex for animal studies; demographics for clinical studies), details of the TBS protocol (e.g., stimulation parameters and target site), timing of assessments, and the outcomes measured.

Given the heterogeneity across study designs, stimulation protocols, and outcome measures, performing a meta-analysis was not feasible.

### 2.4. Risk of bias assessment

To assess the methodological quality and risk of bias in the included studies, we employed the Joanna Briggs Institute (JBI) critical appraisal checklists, tailored to the specific study designs. Each study was independently evaluated by two reviewers using the relevant JBI tool. For each checklist item, responses were rated as “*Yes*,” “*No*,” “*Unclear*,” or “*Not Applicable*.” An overall quality rating (e.g., low, moderate, or high risk of bias) was then determined based on the number and significance of criteria met (23, 24). The quality of the included preclinical studies was evaluated using the Systematic Review Centre for Laboratory Animal Experimentation (SYRCLE) (25), adapted from the Cochrane Collaboration Risk of Bias Tool (26).

## 3. Results

A total of 5,731 articles were initially identified through the database search. Following title and abstract screening, 40 articles were selected for full-text review. Ultimately, 20 journal articles, comprising six preclinical studies (27–32) and 14 clinical studies (33–46) met the inclusion criteria and were included in the systematic review (Figure 1). The overall quality of the included studies was assessed as fair (Table 1).

**Figure 1.**
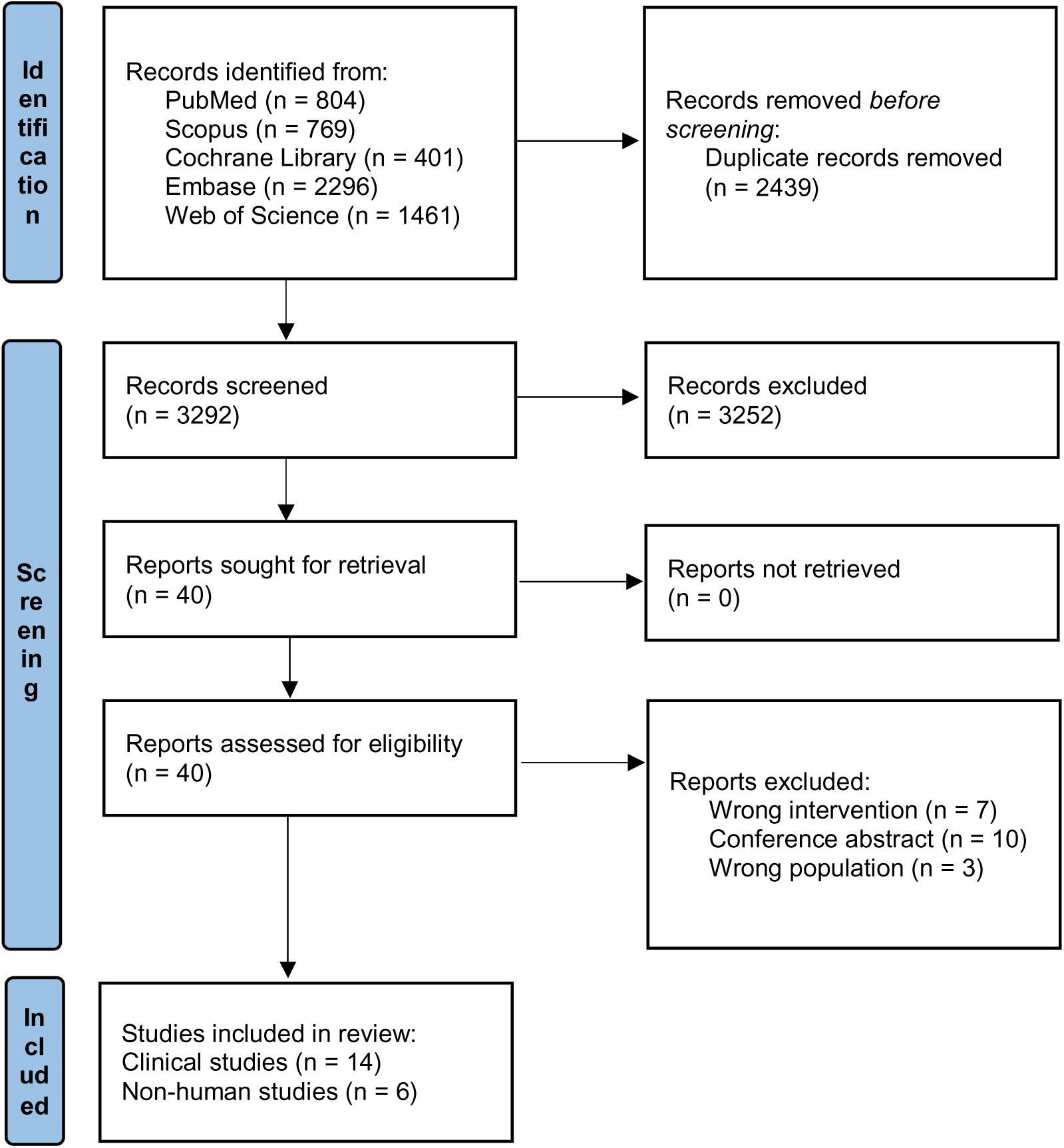
Flow diagram of the included studies

**Table 1.**
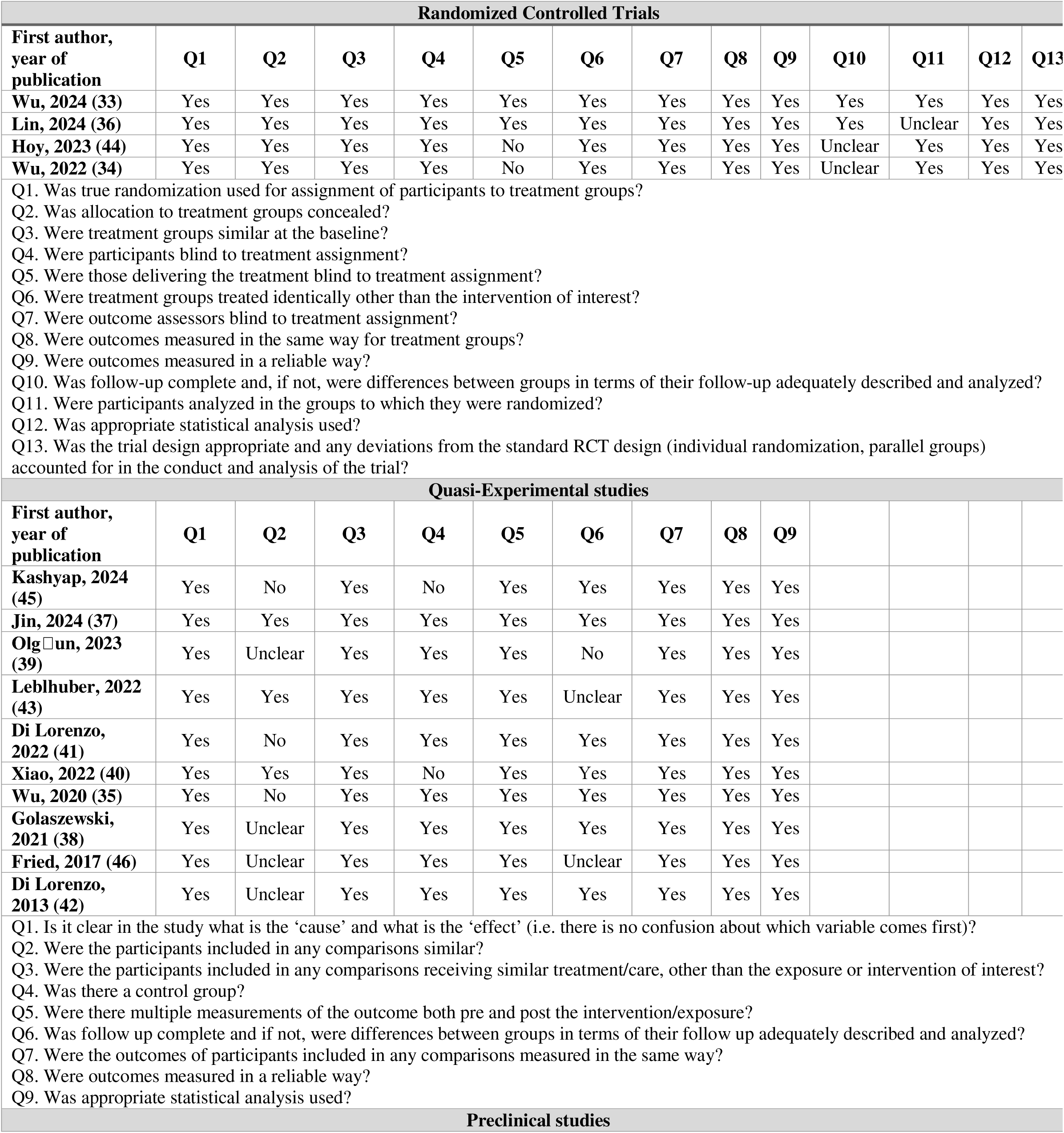
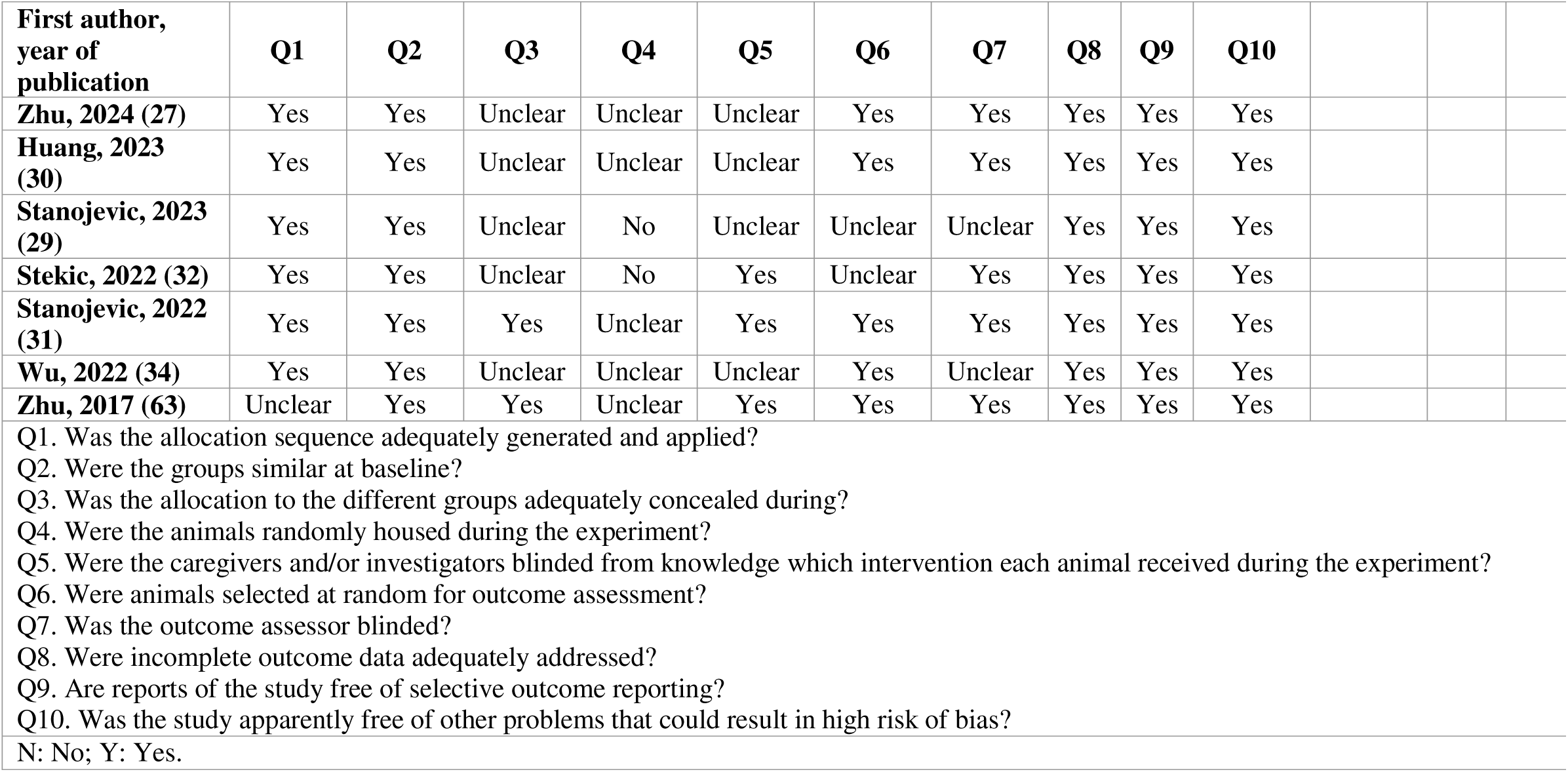
Quality assessment of the included studies.

### 3.1. Preclinical studies

A detailed summary of the animal models, TBS protocols, and assessment time points used in the six included studies is provided in Table 2 (27–32).

**Table 2.** Overview of Preclinical Studies Investigating TBS as a Therapeutic Intervention for Alzheimer’s Disease.

| Study | Model | TBS protocol | Time of Assessment | Outcomes |
| --- | --- | --- | --- | --- |
| **iTBS** |  |  |  |  |
| **Zhu, 2024 (27)** | **6-month-old female APP/PS1 mice and matched wild-type mice.**<br>Divided into three groups: wild-type+Placebo<br>APP/PS1+Placebo<br>APP/PS1+iTBS<br>**Cell culture:** SH-SY5Y cells transfected with the human APP695 gene carrying the Swedish mutant (SH-SY5Y-APP695) | **iTBS group:** Once daily, for 21 consecutive days<br>Intensity: 80-100% MT<br>Number of pulses: in each burst = 3, in total = 1200/session<br>Frequency: intra-burst = 50 Hz, inter-burst = 5 Hz<br>Coil diameter: 3 cm<br>**Placebo groups:** fixed and exposure to noise.<br>**In vitro:** iTBS was administered 24 h after transfection with 600 stimuli per day at the maximum output intensity of the device for five consecutive days. | After TBS treatment. | Improved spatial learning and memory (Morris water maze).<br>Enhanced locomotor and exploratory activities (Open field and Y-maze tests)<br>Reduced Aβ load in the hippocampus and neocortex<br>Attenuated synaptic loss and neuronal apoptosis<br>Increased synaptic density and plasticity<br>Reduced neuroinflammation and oxidative stress markers.<br>Improved glucose uptake<br>Enhanced mitochondrial respiration, fusion, and preserved membrane potential<br>Increased neuronal excitability and activity<br>Upregulation of ISCA1 and ISC assembly proteins<br>Attenuated oxidative damage in the brain<br>No significant difference in swimming speed (Morris water maze), levels of synaptic proteins PSD95 and SNAP25, astrocyte activation in hippocampus, levels of mitochondrial proteins (ACO2, SDHB, COX IV), and expression of mitochondrial fission proteins (DRP1, MFF, FIS1) |
| **Huang, 2023 (30)** | **6-month-old female APP/PS1 mice (N=14).**<br>Divided into two groups: APP/PS1+Placebo (N=7)<br>APP/PS1+iTBS (N=7). | **iTBS group:** Once daily, for 30 consecutive days, after a 7-day adaptation period<br>Intensity: 80% MT<br>Number of pulses: in each burst = 3, in total = 1200/session<br>Frequency: intra-burst = 50 Hz, inter-burst = 5 Hz<br>Duration: 400s/session<br>Coil diameter: 60 mm<br>**Placebo groups:** fixed and exposure to noise. | 2 months after the last iTBS treatment. | Reduced Aβ plaque deposition in the cortex and hippocampus<br>Decreased soluble, diffuse, and fibrillar forms of Aβ<br>Reduced neuronal apoptosis<br>Reduced neuroinflammation (decreased GFAP-positive astrocytes, Iba1-positive, and CD68-positive microglia)<br>Improved synaptic integrity (increased NeuN and MAP-2 fluorescence; increased PSD95, SYN1, SNAP25)<br>Increased BDNF levels<br>No significant changes in APP levels (full-length and soluble), alterations in ADAM10 enzyme, levels of NEP, LRP1, RAGE, PSD93, and VAMP1 proteins |
| **Stanojevic, 2023 (29)** | **10-weeks-old male Wistar rats (N=35).** | **iTBS group:** Once daily, for two 5-day sessions with a two-day pause in between, starting 5 weeks after STZ | After TBS treatment. | Reduced oxidative and nitrosative stress (decreased superoxide anion, MDA, NO□+NO□)<br>Attenuation of nucleic acid damage markers (8OHdG and EGR1) |
|  | Divided into four groups:<br>Control (N=9)<br>STZ (N=9)<br>STZ+iTBS (N=9)<br>STZ+Placebo (N=8)<br>All animals underwent stereotaxic surgery. STZ groups received a bilateral icv injection of streptozotocin solution (3 mg/kg).<br>Control groups received icv of saline solution. | injection<br>1) Intensity: 33% of maximum output<br>2) Number of pulses: in each burst = 3, in total = 600/session<br>3) Frequency: intra-burst = 50 Hz, inter-burst = 5 Hz<br>4) Duration: 2s, with 8s intervals, for 192s/session<br>5) Coil diameter: 25 mm, figure-of-eight<br>• **Placebo groups:** manual manipulation and exposure to noise |  | • Reduced APP in cortex and striatum, and reduced Aβ in all tested regions (cortex, striatum, hippocampus, cerebellum)<br>• Enhanced enzymatic antioxidants (increased tSOD, CuZnSOD, MnSOD, CAT activity)<br>• Improved non-enzymatic antioxidants (increased SH in all regions, increased GSH specifically in hippocampus and cerebellum)<br>• Elevated Nrf2 and BDNF levels<br>• Reduced reactive astrogliosis in hippocampus (decreased GFAP/VIM/C3 co-localization)<br>• No significant changes in APP levels in hippocampus and cerebellum and hippocampal Aβ levels according to dot-blot analysis (contrasting with ELISA findings) |
| **Stanojevic, 2022 (31)** | **10-weeks-old male Wistar rats (N=44).**<br>Divided into five groups:<br>Control<br>STZ<br>STZ+iTBS<br>STZ+Placebo<br>Control+iTBS<br>All animals underwent stereotaxic surgery. STZ groups received a bilateral icv injection of streptozotocin solution (3 mg/kg).<br>Control groups received icv of saline solution. | • **iTBS group:** Once daily, for two 5-day sessions with a two-day pause in between, starting 37 days after STZ injection<br>1) Intensity: 33% of maximum output<br>2) Number of pulses: in each burst = 3, in total = 600/session<br>3) Frequency: intra-burst = 50 Hz, inter-burst = 5 Hz<br>4) Duration: 2s, with 8s intervals, for 192s/session<br>5) Coil diameter: 25 mm, figure-of-eight<br>• **Placebo groups:** manual manipulation and exposure to noise | After TBS treatment | • Improved cognitive impairment (reduced task execution time and improved memory scores in eight arm radial maze tests)<br>• Reduced reactive gliosis: decreased microglial activation in dorsal hippocampus, medial habenula, prefrontal cortex, and caudoputamen<br>• Reduced reactive astrogliosis: decreased GFAP+/VIM+ expression in hippocampal regions (CA1 and fimbria) and periventricular caudoputamen<br>• No significant motor deficits induced by STZ administration<br>• Mild histomorphological changes persisted in ventral hippocampus, medial habenula, and periventricular regions after iTBS treatment |
| **Stekic, 2022 (32)** | **2-months-old, male Wistar rats (N=54)**<br>Divided into four groups:<br>Control (N=12)<br>TMT (N=15)<br>TMT+iTBS (N=12)<br>TMT+placebo (N=15) | • **iTBS group:** Twice daily, for 15 days, starting 3 days after TMT injection<br>1) Intensity: 33% of maximum output<br>2) Number of pulses: in each burst = 3, in total = 600/session<br>3) Frequency: intra-burst = 50 Hz, inter-burst = 5 Hz | 3 weeks after TMT injection, with iTBS starting 3 days after TMT. | • Improved cognitive performance (Novel Object Recognition Test and Open Field Test)<br>• Reduced hyperactivity, aggressive behavior, tremors, and seizures induced by TMT<br>• Reduced neuronal apoptosis (reduced Bax/Bcl-2 ratio)<br>• Reduced hippocampal neuronal loss and preservation of hippocampal structure<br>• Attenuated neuroinflammation (reduced astrocyte activation, decreased IL-1β) |
|  | TMT groups received a single intraperitoneal injection of Trimethyltin (8 mg/kg).<br>Control group received a single intraperitoneal injection of 0.9% saline solution. | 4) Duration: 2s, with 8s intervals, for 192s/session<br>5) Coil diameter: 25 mm, figure-of-eight<br>**Placebo groups:** manual manipulation and exposure to noise | Daily behavioral scoring for 3 weeks (aggressive behavior). | and complement C3 levels)<br>Enhanced anti-inflammatory response (increased IL-10 expression)<br>Reduced oxidative and nitrosative stress (restored activity of total SOD, MnSOD, decreased levels of O<sub>2</sub><sup>-</sup>, MDA, and NO<sub>2</sub><sup>-</sup>)<br>Improved antioxidative status (increased SH<sub>2</sub> and total GSH content)<br>Restored PI3K/Akt/mTOR and ERK1/2 signaling pathway activity<br>No significant recovery in CuZnSOD enzyme activity after iTBS treatment<br>Astrocytic activation persisted despite reduced inflammatory markers (astrocytes maintained a reactive phenotype) |

| cTBS |  |  |  |  |
| --- | --- | --- | --- | --- |
| **Wu, 2022 (28)** | **7-8-months-old male APP/PS1 mice (N=12)**<br>Divided into two groups:<br>cTBS (N=6)<br>Placebo (N=6) | **cTBS group:** once daily, for 4 weeks<br>Intensity: 25% of maximum output<br>Number of pulses: 2000/session<br>Frequency: intra-burst = 50 Hz, inter-burst = 5 Hz<br>Duration: A 40s pulse train<br>Coil diameter: 70 mm<br>**Placebo groups:** fixed and exposure to noise. | immediately after<br>two-photon imaging 1 week after<br>brain tissue collection 9 days after cTBS treatment. | Improved spatial cognitive function (Morris water maze)<br>Reduced Aβ deposition in cortex and hippocampus<br>Improved polarization (perivascular localization) of aquaporin-4<br>Significant correlation between enhanced glymphatic clearance, reduced Aβ deposition, and improved cognitive function<br>Improved astrocytic aquaporin-4 polarization<br>No significant change in total expression levels of AQP4 in cortex or hippocampus |

Animal studies investigating the effects of TBS on AD have utilized different rodent models, including transgenic APP/PS1 mice (27, 28, 30) and rats with streptozotocin (STZ)-induced (29, 31) or trimethyltin (TMT)-induced neurodegeneration (32), replicating key pathological features of AD, such as Aβ accumulation, neuroinflammation, oxidative stress, and cognitive impairment (27–32). The included studies applied TBS using different stimulation paradigms, varying in the number of trains, pulses per burst, and inter-train intervals. Only one study used cTBS while the other applied iTBS. Stimulation was administered either once daily (28–31) or twice daily (32), over treatment durations ranging from 10 days to one month, with some protocols incorporating breaks between sessions. Stimulation intensities ranged from 33% to 100% of the motor thresholds (MTs). Different coil configurations, including figure-of-eight (25 mm) and circular (60 mm) coils, were employed. Placebo conditions involved noise exposure and manual manipulation without active stimulation.

The time of assessment varied across studies, ranging from immediately after TBS treatment (27–29) to follow-up periods of two months post-treatment (30). One study assessed behavioral outcomes during stimulation periods (32), while others evaluated long-term effects after the final TBS session. Studies consistently demonstrate that TBS exerts neuroprotective effects in AD models by reducing pathological hallmarks, enhancing synaptic plasticity, and improving cognitive function.

TBS reduced Aβ burden by downregulating *BACE1* (inhibition of Aβ production) and upregulating *IDE* (facilitation of Aβ degradation) (27, 29, 30). TBS also promoted glymphatic clearance and improved astrocytic aquaporin-4 (*AQP4*) polarization, the processes that were correlated with enhanced Aβ removal and cognitive improvements (28).

TBS mitigated neuronal apoptosis, as indicated by decreased expression of pro-apoptotic markers (*Caspase-3*, *Bax*) and increased levels of anti-apoptotic and neuronal survival markers (*Bcl-2*, *NeuN*) (27, 30, 32). Synaptic integrity was preserved through increased expression of synaptic proteins, including *PSD95*, *SYN1*, *SNAP25*, and *VAMP1*, while upregulation of *BDNF* further supported neuroplasticity (27, 29, 30). Electrophysiological assessments also revealed that TBS enhanced neuronal excitability and reduced hypoactivity in cortical and hippocampal neurons (27).

In addition to neuroprotection, TBS exhibited potent anti-inflammatory and antioxidative effects. Studies reported reductions in microglial and astrocytic activation, as evidenced by decreased expression of *Iba1*, *CD68*, and *GFAP*, along with lower levels of pro-inflammatory cytokines (*IL-1*β, *IL-6*, *TNF-*α, *IFN-*γ) (27–32). TBS also restored oxidative balance by enhancing antioxidant enzyme activity (*tSOD, MnSOD, GSH*) and reducing oxidative stress markers (*MDA, 3-NT, NO2−*) (32). Furthermore, TBS modulated key signaling pathways involved in cellular survival and synaptic function, including *PI3K/Akt/mTOR* and *ERK1/2*, which were restored following stimulation (32).

### 3.2. Clinical studies

An overview of the 14 clinical studies included in this review is provided in Table 3 (33–46). Collectively, these studies included 324 patients with AD, and 269 controls.

**Table 3.**
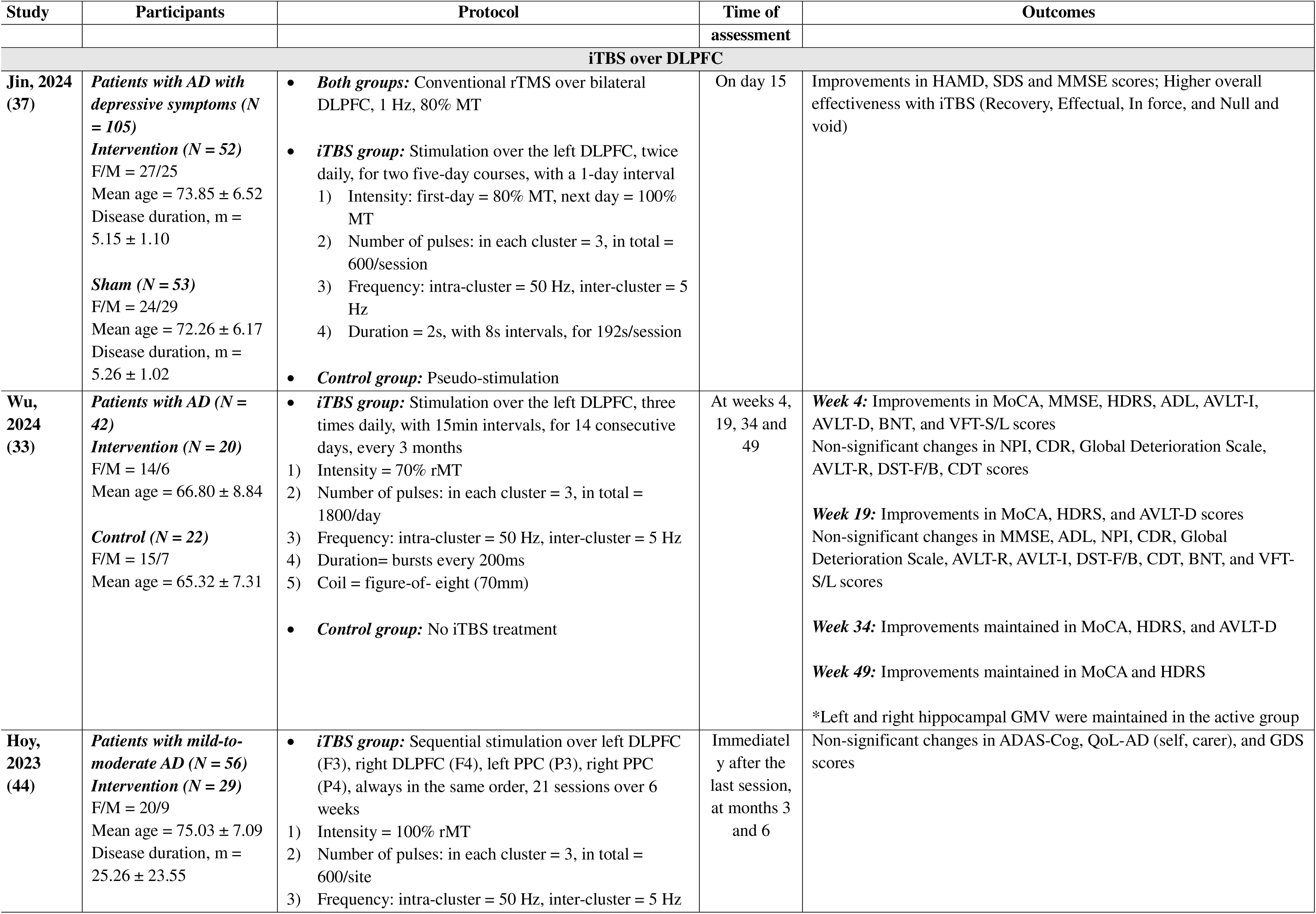
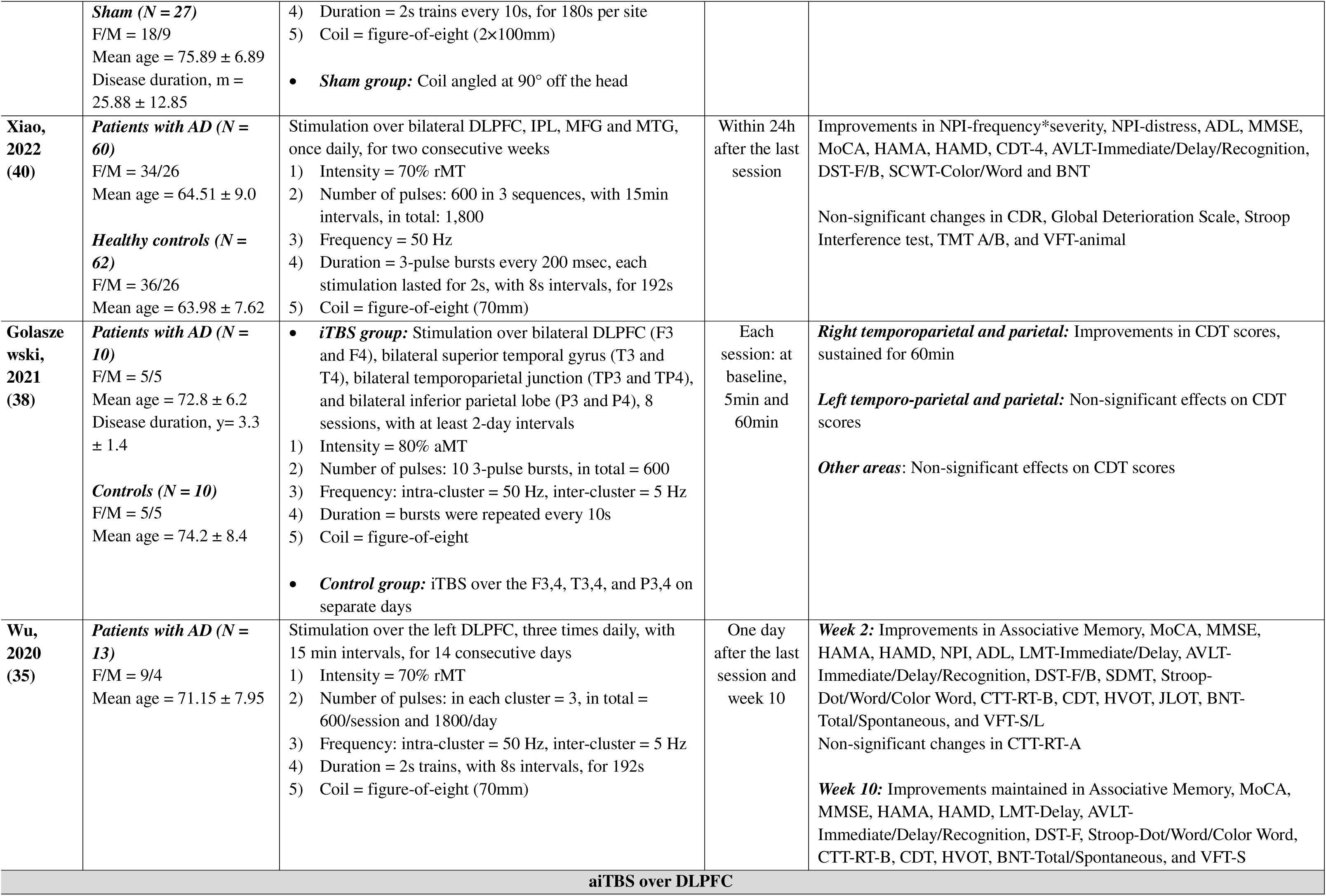
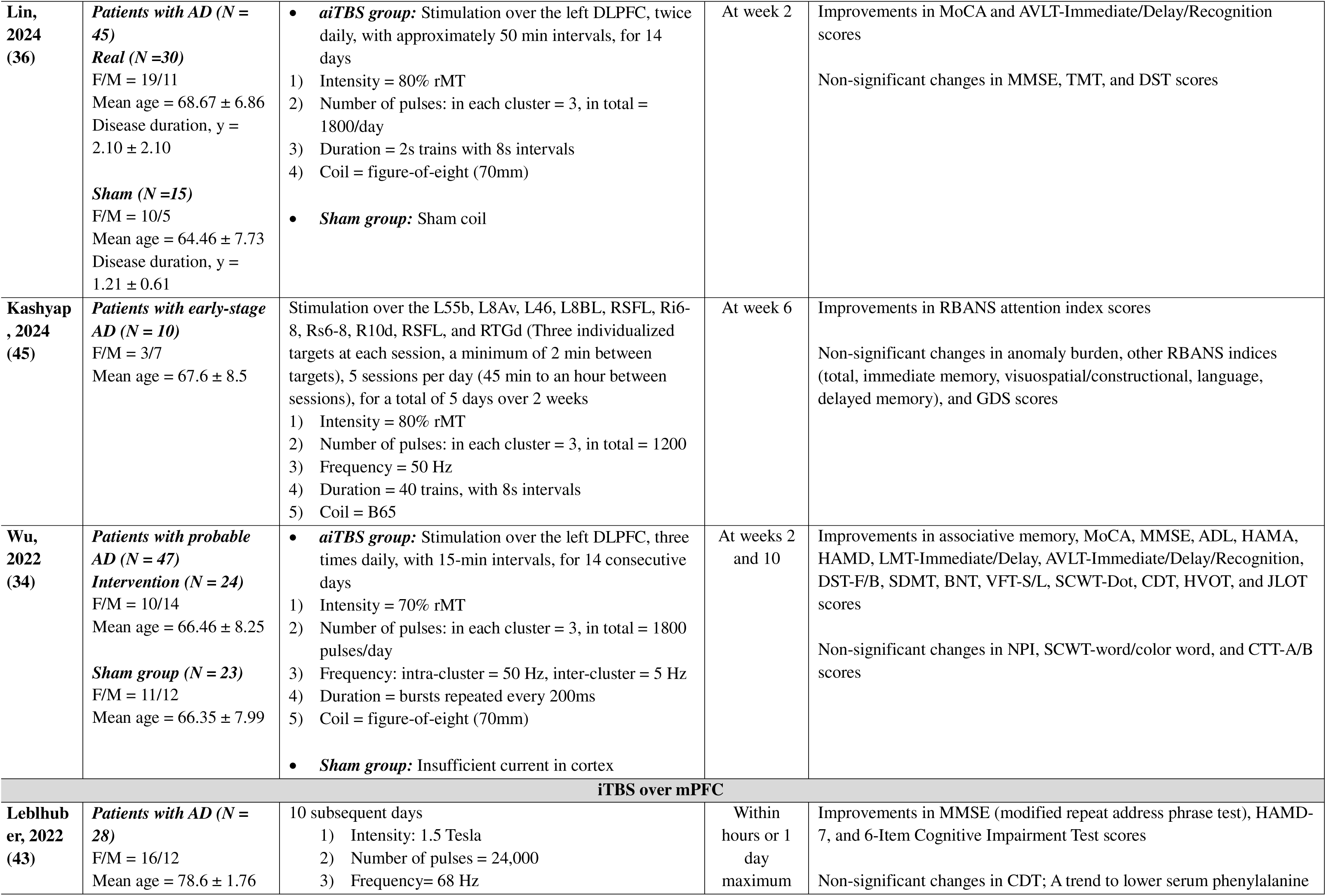
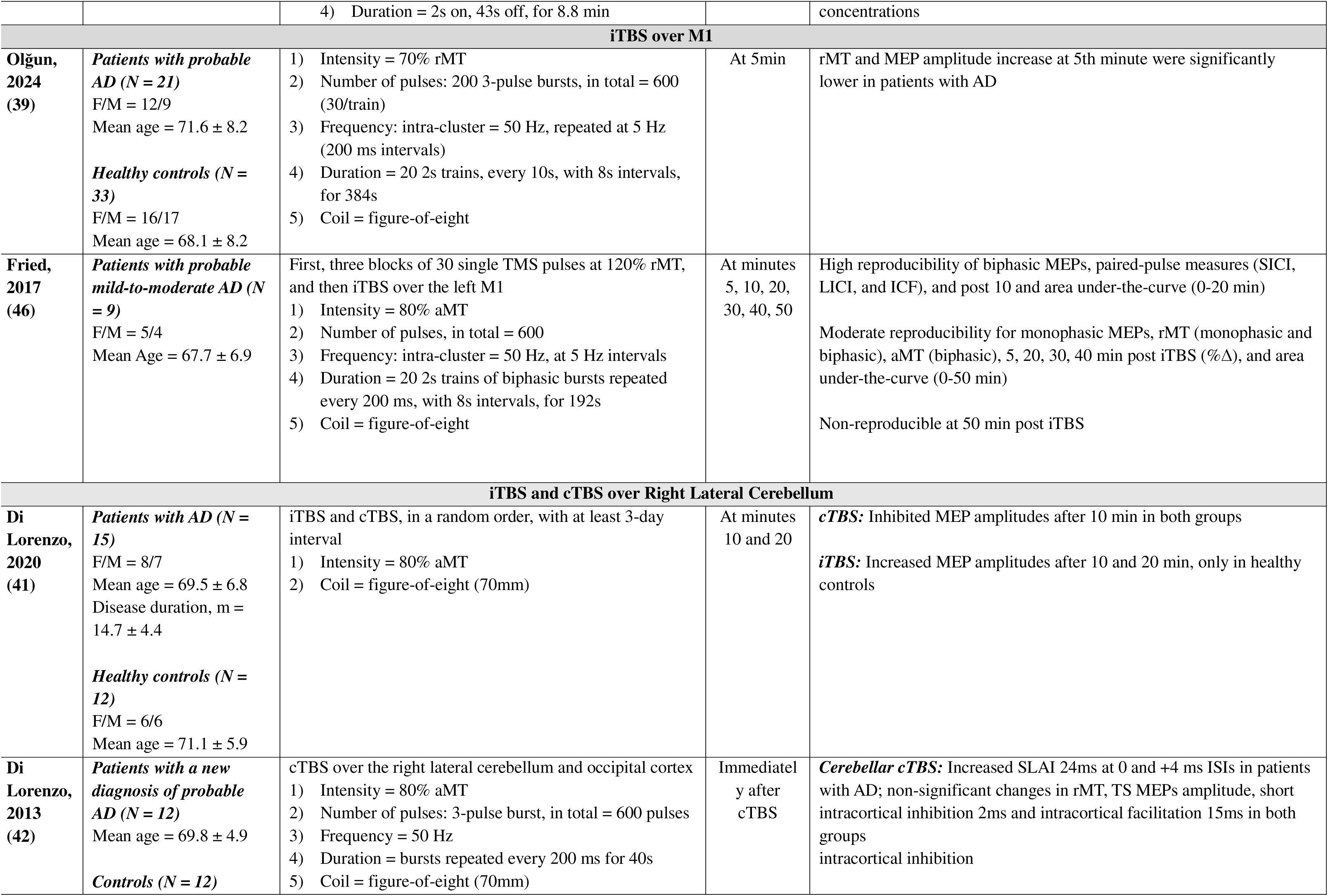
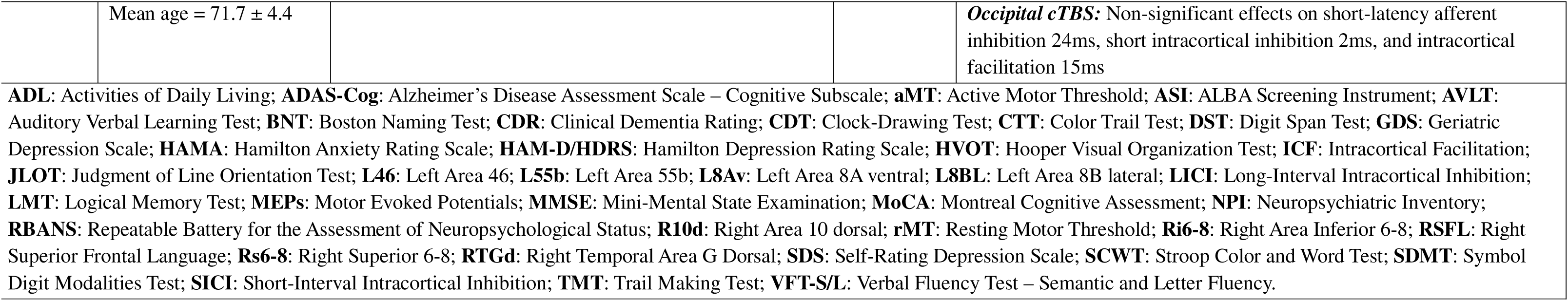
Overview of Clinical Studies Investigating TBS as a Therapeutic Intervention for Alzheimer’s Disease.

| Study | Participants | Protocol | Time of | Outcomes |
| --- | --- | --- | --- | --- |

|  |  |  | assessment |  |
| --- | --- | --- | --- | --- |
| **iTBS over DLPFC** |  |  |  |  |
| **Jin, 2024**<br>**(37)** | **Patients with AD with depressive symptoms** (*N* = 105)<br>**Intervention** (*N* = 52)<br>F/M = 27/25<br>Mean age = 73.85 ± 6.52<br>Disease duration, m = 5.15 ± 1.10<br>**Sham** (*N* = 53)<br>F/M = 24/29<br>Mean age = 72.26 ± 6.17<br>Disease duration, m = 5.26 ± 1.02 | **Both groups:** Conventional rTMS over bilateral DLPFC, 1 Hz, 80% MT<br>**iTBS group:** Stimulation over the left DLPFC, twice daily, for two five-day courses, with a 1-day interval<br>Intensity: first-day = 80% MT, next day = 100% MT<br>Number of pulses: in each cluster = 3, in total = 600/session<br>Frequency: intra-cluster = 50 Hz, inter-cluster = 5 Hz<br>Duration = 2s, with 8s intervals, for 192s/session<br>**Control group:** Pseudo-stimulation | On day 15 | Improvements in HAMD, SDS and MMSE scores; Higher overall effectiveness with iTBS (Recovery, Effectual, In force, and Null and void) |
| **Wu, 2024**<br>**(33)** | **Patients with AD** (*N* = 42)<br>**Intervention** (*N* = 20)<br>F/M = 14/6<br>Mean age = 66.80 ± 8.84<br>**Control** (*N* = 22)<br>F/M = 15/7<br>Mean age = 65.32 ± 7.31 | **iTBS group:** Stimulation over the left DLPFC, three times daily, with 15min intervals, for 14 consecutive days, every 3 months<br>Intensity = 70% rMT<br>Number of pulses: in each cluster = 3, in total = 1800/day<br>Frequency: intra-cluster = 50 Hz, inter-cluster = 5 Hz<br>Duration= bursts every 200ms<br>Coil = figure-of- eight (70mm)<br>**Control group:** No iTBS treatment | At weeks 4, 19, 34 and 49 | **Week 4:** Improvements in MoCA, MMSE, HDRS, ADL, AVLT-I, AVLT-D, BNT, and VFT-S/L scores<br>Non-significant changes in NPI, CDR, Global Deterioration Scale, AVLT-R, DST-F/B, CDT scores<br>**Week 19:** Improvements in MoCA, HDRS, and AVLT-D scores<br>Non-significant changes in MMSE, ADL, NPI, CDR, Global Deterioration Scale, AVLT-R, AVLT-I, DST-F/B, CDT, BNT, and VFT-S/L scores<br>**Week 34:** Improvements maintained in MoCA, HDRS, and AVLT-D<br>**Week 49:** Improvements maintained in MoCA and HDRS<br>*Left and right hippocampal GMV were maintained in the active group |
| **Hoy, 2023**<br>**(44)** | **Patients with mild-to-moderate AD** (*N* = 56)<br>**Intervention** (*N* = 29)<br>F/M = 20/9<br>Mean age = 75.03 ± 7.09<br>Disease duration, m = 25.26 ± 23.55 | **iTBS group:** Sequential stimulation over left DLPFC (F3), right DLPFC (F4), left PPC (P3), right PPC (P4), always in the same order, 21 sessions over 6 weeks<br>Intensity = 100% rMT<br>Number of pulses: in each cluster = 3, in total = 600/site<br>Frequency: intra-cluster = 50 Hz, inter-cluster = 5 Hz | Immediately after the last session, at months 3 and 6 | Non-significant changes in ADAS-Cog, QoL-AD (self, carer), and GDS scores |
|  | **Sham** (*N* = 27)<br>F/M = 18/9<br>Mean age = 75.89 ± 6.89<br>Disease duration, m = 25.88 ± 12.85 | 4) Duration = 2s trains every 10s, for 180s per site<br>5) Coil = figure-of-eight (2×100mm)<br>• **Sham group:** Coil angled at 90° off the head |  |  |
| **Xiao, 2022 (40)** | **Patients with AD** (*N* = 60)<br>F/M = 34/26<br>Mean age = 64.51 ± 9.0<br>**Healthy controls** (*N* = 62)<br>F/M = 36/26<br>Mean age = 63.98 ± 7.62 | Stimulation over bilateral DLPFC, IPL, MFG and MTG, once daily, for two consecutive weeks<br>1) Intensity = 70% rMT<br>2) Number of pulses: 600 in 3 sequences, with 15min intervals, in total: 1,800<br>3) Frequency = 50 Hz<br>4) Duration = 3-pulse bursts every 200 msec, each stimulation lasted for 2s, with 8s intervals, for 192s<br>5) Coil = figure-of-eight (70mm) | Within 24h after the last session | Improvements in NPI-frequency*severity, NPI-distress, ADL, MMSE, MoCA, HAMA, HAMD, CDT-4, AVLT-Immediate/Delay/Recognition, DST-F/B, SCWT-Color/Word and BNT<br>Non-significant changes in CDR, Global Deterioration Scale, Stroop Interference test, TMT A/B, and VFT-animal |
| **Golaszewski, 2021 (38)** | **Patients with AD** (*N* = 10)<br>F/M = 5/5<br>Mean age = 72.8 ± 6.2<br>Disease duration, y = 3.3 ± 1.4<br>**Controls** (*N* = 10)<br>F/M = 5/5<br>Mean age = 74.2 ± 8.4 | • **iTBS group:** Stimulation over bilateral DLPFC (F3 and F4), bilateral superior temporal gyrus (T3 and T4), bilateral temporoparietal junction (TP3 and TP4), and bilateral inferior parietal lobe (P3 and P4), 8 sessions, with at least 2-day intervals<br>1) Intensity = 80% aMT<br>2) Number of pulses: 10 3-pulse bursts, in total = 600<br>3) Frequency: intra-cluster = 50 Hz, inter-cluster = 5 Hz<br>4) Duration = bursts were repeated every 10s<br>5) Coil = figure-of-eight<br>• **Control group:** iTBS over the F3,4, T3,4, and P3,4 on separate days | Each session: at baseline, 5min and 60min | **Right temporoparietal and parietal:** Improvements in CDT scores, sustained for 60min<br>**Left temporo-parietal and parietal:** Non-significant effects on CDT scores<br>**Other areas:** Non-significant effects on CDT scores |
| **Wu, 2020 (35)** | **Patients with AD** (*N* = 13)<br>F/M = 9/4<br>Mean age = 71.15 ± 7.95 | Stimulation over the left DLPFC, three times daily, with 15 min intervals, for 14 consecutive days<br>1) Intensity = 70% rMT<br>2) Number of pulses: in each cluster = 3, in total = 600/session and 1800/day<br>3) Frequency: intra-cluster = 50 Hz, inter-cluster = 5 Hz<br>4) Duration = 2s trains, with 8s intervals, for 192s<br>5) Coil = figure-of-eight (70mm) | One day after the last session and week 10 | **Week 2:** Improvements in Associative Memory, MoCA, MMSE, HAMA, HAMD, NPI, ADL, LMT-Immediate/Delay, AVLT-Immediate/Delay/Recognition, DST-F/B, SDMT, Stroop-Dot/Word/Color Word, CTT-RT-B, CDT, HVOT, JLOT, BNT-Total/Spontaneous, and VFT-S/L<br>Non-significant changes in CTT-RT-A<br>**Week 10:** Improvements maintained in Associative Memory, MoCA, MMSE, HAMA, HAMD, LMT-Delay, AVLT-Immediate/Delay/Recognition, DST-F, Stroop-Dot/Word/Color Word, CTT-RT-B, CDT, HVOT, BNT-Total/Spontaneous, and VFT-S |
| **aiTBS over DLPFC** |  |  |  |  |
| **Lin, 2024 (36)** | **Patients with AD (N = 45)**<br>**Real (N = 30)**<br>F/M = 19/11<br>Mean age = 68.67 ± 6.86<br>Disease duration, y = 2.10 ± 2.10<br>**Sham (N = 15)**<br>F/M = 10/5<br>Mean age = 64.46 ± 7.73<br>Disease duration, y = 1.21 ± 0.61 | **aiTBS group:** Stimulation over the left DLPFC, twice daily, with approximately 50 min intervals, for 14 days<br>Intensity = 80% rMT<br>Number of pulses: in each cluster = 3, in total = 1800/day<br>Duration = 2s trains with 8s intervals<br>Coil = figure-of-eight (70mm)<br>**Sham group:** Sham coil | At week 2 | Improvements in MoCA and AVLT-Immediate/Delay/Recognition scores<br>Non-significant changes in MMSE, TMT, and DST scores |
| **Kashyap, 2024 (45)** | **Patients with early-stage AD (N = 10)**<br>F/M = 3/7<br>Mean age = 67.6 ± 8.5 | Stimulation over the L55b, L8Av, L46, L8BL, RSFL, Ri6-8, Rs6-8, R10d, RSFL, and RTGd (Three individualized targets at each session, a minimum of 2 min between targets), 5 sessions per day (45 min to an hour between sessions), for a total of 5 days over 2 weeks<br>Intensity = 80% rMT<br>Number of pulses: in each cluster = 3, in total = 1200<br>Frequency = 50 Hz<br>Duration = 40 trains, with 8s intervals<br>Coil = B65 | At week 6 | Improvements in RBANS attention index scores<br>Non-significant changes in anomaly burden, other RBANS indices (total, immediate memory, visuospatial/constructional, language, delayed memory), and GDS scores |
| **Wu, 2022 (34)** | **Patients with probable AD (N = 47)**<br>**Intervention (N = 24)**<br>F/M = 10/14<br>Mean age = 66.46 ± 8.25<br>**Sham group (N = 23)**<br>F/M = 11/12<br>Mean age = 66.35 ± 7.99 | **aiTBS group:** Stimulation over the left DLPFC, three times daily, with 15-min intervals, for 14 consecutive days<br>Intensity = 70% rMT<br>Number of pulses: in each cluster = 3, in total = 1800 pulses/day<br>Frequency: intra-cluster = 50 Hz, inter-cluster = 5 Hz<br>Duration = bursts repeated every 200ms<br>Coil = figure-of-eight (70mm)<br>**Sham group:** Insufficient current in cortex | At weeks 2 and 10 | Improvements in associative memory, MoCA, MMSE, ADL, HAMA, HAMD, LMT-Immediate/Delay, AVLT-Immediate/Delay/Recognition, DST-F/B, SDMT, BNT, VFT-S/L, SCWT-Dot, CDT, HVOT, and JLOT scores<br>Non-significant changes in NPI, SCWT-word/color word, and CTT-A/B scores |
| **iTBS over mPFC** |  |  |  |  |
| **Leblhuber, 2022 (43)** | **Patients with AD (N = 28)**<br>F/M = 16/12<br>Mean age = 78.6 ± 1.76 | 10 subsequent days<br>Intensity: 1.5 Tesla<br>Number of pulses = 24,000<br>Frequency= 68 Hz | Within hours or 1 day maximum | Improvements in MMSE (modified repeat address phrase test), HAMD-7, and 6-Item Cognitive Impairment Test scores<br>Non-significant changes in CDT; A trend to lower serum phenylalanine |
|  |  | 4) Duration = 2s on, 43s off, for 8.8 min |  | concentrations |
| **iTBS over M1** |  |  |  |  |
| **Olğun, 2024 (39)** | *Patients with probable AD (N = 21)*<br>F/M = 12/9<br>Mean age = 71.6 ± 8.2<br>*Healthy controls (N = 33)*<br>F/M = 16/17<br>Mean age = 68.1 ± 8.2 | 1) Intensity = 70% rMT<br>2) Number of pulses: 200 3-pulse bursts, in total = 600 (30/train)<br>3) Frequency: intra-cluster = 50 Hz, repeated at 5 Hz (200 ms intervals)<br>4) Duration = 20 2s trains, every 10s, with 8s intervals, for 384s<br>5) Coil = figure-of-eight | At 5min | rMT and MEP amplitude increase at 5th minute were significantly lower in patients with AD |
| **Fried, 2017 (46)** | *Patients with probable mild-to-moderate AD (N = 9)*<br>F/M = 5/4<br>Mean Age = 67.7 ± 6.9 | First, three blocks of 30 single TMS pulses at 120% rMT, and then iTBS over the left M1<br>1) Intensity = 80% aMT<br>2) Number of pulses, in total = 600<br>3) Frequency: intra-cluster = 50 Hz, at 5 Hz intervals<br>4) Duration = 20 2s trains of biphasic bursts repeated every 200 ms, with 8s intervals, for 192s<br>5) Coil = figure-of-eight | At minutes 5, 10, 20, 30, 40, 50 | High reproducibility of biphasic MEPs, paired-pulse measures (SICI, LICI, and ICF), and post 10 and area under-the-curve (0-20 min)<br>Moderate reproducibility for monophasic MEPs, rMT (monophasic and biphasic), aMT (biphasic), 5, 20, 30, 40 min post iTBS (%Δ), and area under-the-curve (0-50 min)<br>Non-reproducible at 50 min post iTBS |
| **iTBS and cTBS over Right Lateral Cerebellum** |  |  |  |  |
| **Di Lorenzo, 2020 (41)** | *Patients with AD (N = 15)*<br>F/M = 8/7<br>Mean age = 69.5 ± 6.8<br>Disease duration, m = 14.7 ± 4.4<br>*Healthy controls (N = 12)*<br>F/M = 6/6<br>Mean age = 71.1 ± 5.9 | iTBS and cTBS, in a random order, with at least 3-day interval<br>1) Intensity = 80% aMT<br>2) Coil = figure-of-eight (70mm) | At minutes 10 and 20 | *cTBS*: Inhibited MEP amplitudes after 10 min in both groups<br>*iTBS*: Increased MEP amplitudes after 10 and 20 min, only in healthy controls |
| **Di Lorenzo, 2013 (42)** | *Patients with a new diagnosis of probable AD (N = 12)*<br>Mean age = 69.8 ± 4.9<br>*Controls (N = 12)* | cTBS over the right lateral cerebellum and occipital cortex<br>1) Intensity = 80% aMT<br>2) Number of pulses: 3-pulse burst, in total = 600 pulses<br>3) Frequency = 50 Hz<br>4) Duration = bursts repeated every 200 ms for 40s<br>5) Coil = figure-of-eight (70mm) | Immediately after cTBS | *Cerebellar cTBS*: Increased SLAI 24ms at 0 and +4 ms ISIs in patients with AD; non-significant changes in rMT, TS MEPs amplitude, short intracortical inhibition 2ms and intracortical facilitation 15ms in both groups intracortical inhibition |
|  | Mean age = 71.7 ± 4.4 |  |  | ***Occipital cTBS:*** Non-significant effects on short-latency afferent inhibition 24ms, short intracortical inhibition 2ms, and intracortical facilitation 15ms |

TBS protocols varied in terms of stimulation targets, frequency, duration, and assessment time points. Most studies applied iTBS or accelerated iTBS (aiTBS) over the left DLPFC (33, 36–38, 40, 44), with others targeting other brain regions such as the primary motor cortex (39), medial prefrontal cortex (43), and cerebellum (41, 42). Stimulation intensity ranged from 70% to 100% of the motor threshold (MT), with total pulses varying from 600 and 24,000. Protocols typically involved daily or multiple daily sessions over a period of 5 to 14 days, with follow-up assessments ranging from immediately post-treatment to several months. Sham protocols consistently mimicked auditory and tactile sensations without delivering active stimulation.

#### 3.2.1. DLPFC

iTBS was found to significantly improve cognitive function as measured by MoCA (33, 35, 36, 40), MMSE (33, 35, 37, 40), associative memory (35), CDT (35, 38, 40), VFT-S/L (33, 35), BNT (33, 35, 40), DST (35, 40), AVLT (33, 35, 36, 40), SDMT, CTT-RT-B, HVOT, JLOT, LMT (35), and SCWT (40), depression as measured by SDS (37) and HAMD (33, 35, 37, 40), anxiety assessed through HAMA (35, 40), ADL (33, 35, 40), NPI (35, 40), and overall treatment effectiveness (recovery, effectual, in Force, and null and void) (37). However, some studies reported non-significant findings across NPI (total score, caregiver distress), CTT-RT-A (35), Global Deterioration Scale (33, 40), CDR, Stroop Interference test, TMT A/B, and VFT-animal (40), Geriatric Depression Scale, ADAS-Cog, and quality of life (self, carer) (44).

aiTBS significantly improved associative memory, MMSE, ADL, HAMA, HAMD, LMT-Immediate/Delay, DST-F/B, SDMT, BNT, VFT-S/L, SCWT-Dot, CDT, HVOT, and JLOT scores (34), MoCA, AVLT-Immediate/Delay/Recognition scores (34, 36), and the RBANS attention index (45). Also, following treatment, there was a trend toward the improvement of story memory which is a part of the immediate memory index (45). No significant changes were found in anomaly burden, or other RBANS indices (total, immediate memory, visuospatial/constructional, language, delayed memory) (45). Also, non-significant findings were reported across MMSE, TMT, DST (36), Geriatric Depression Scale (45), NPI, SCWT-word/color word, and CTT-A/B scores (34).

Regarding brain connectivity, iTBS significantly reduced the abnormally elevated connection between the target and right precuneus cortex, which significantly correlated with improvements in VFT-S scores (47). Moreover, the predictors of change in delayed recall post-iTBS were baseline gamma connectivity (i.e., higher gamma connectivity at baseline predicting greater change in delayed recall) and change in gamma connectivity (i.e., greater change in gamma connectivity predicting greater change in delayed recall) (44). Significant changes in global mean field amplitude values (P30 and P70) were found post-iTBS, predominantly located at the left frontal region. Furthermore, total MoCA and MMSE scores were inversely correlated with cortical-evoked activity in the DLPFC. Changes in beta oscillations, also concentrated in the left frontal region, were observed (36).

In terms of gray matter volume (GMV), the left and right hippocampal volumes were preserved in the active treatment group but significantly declined in the sham group. Increases in left and right hippocampal GMV were positively correlated with improvements in MoCA and MMSE scores. Also, left hippocampal GMV increases were negatively correlated with Global Deterioration Scale scores, while right hippocampal GMV increases were positively correlated with VFT-S scores. MoCA scores one year after DLPFC-iTBS treatment significantly improved in non-Apolipoprotein E epsilon 4 (ApoE ε4) carriers, with no such improvements in ApoE ε4 carriers. Improvements in CDT performance were also more pronounced in non-ApoE ε4 carriers, with no significant changes noted in other neuropsychological assessments (33).

#### 3.2.2. M1

Regarding neuroplasticity, the percentage amplitude changes of motor evoked potentials (MEPs) and rMTs were significantly lower in patients with AD, compared to healthy controls, and showed no significant correlation with MMSE, ALBA, CDR, or Global Deterioration Scale scores (39). Another study reported a lower increase in rMT and MEP amplitude at 5 minutes post-iTBS in patients with AD. High reproducibility was observed at 10 minutes post-iTBS, as well as for the total area under the curve during the first 20 minutes following stimulation (AUC –), which corresponds to the typical peak effect period seen in neurotypical individuals. In contrast, only moderate reproducibility was noted at the 5-, 20-, 30-, and 40-minute time points. By 50 minutes post-iTBS, thesed changes were found to be non-reproducible (46).

#### 3.2.3. Medial prefrontal cortex (mPFC)

iTBS over the mPFC significantly improved MMSE (modified repeat address phrase test), 7-item HAMD, and the 6-Item Cognitive Impairment test scores. No significant changes were observed in CDT scores. A trend toward lower serum phenylalanine concentrations was noted (43).

#### 3.2.4. Cerebellum

Two studies targeted the right lateral cerebellum, and subsequently, the posterior and superior lobules. One study indicated a reduction in MEP amplitude 10 minutes after cTBS in both patients and healthy controls, suggesting motor cortex activity suppression. Conversely, iTBS significantly increased MEP amplitude at 10 and 20 minutes post-stimulation, but only in healthy controls (41). Another study found that cTBS significantly increased short-latency afferent inhibition (SLAI) in patients with AD at inter-stimulus intervals (ISIs) of 0 ms and +4 ms, with no effects on rMT, intracortical inhibition (SICI) or intracortical facilitation (ICF) in either group (42).

#### 3.2.5. Adverse events

Overall, the procedure was well-tolerated, with the exception of two participants who could not endure the full stimulation intensity and discontinued treatment (45). Reported adverse events in the active treatment groups included scalp painful discomfort (N = 9) (33, 34, 36, 47), eyelid twitches (N = 5) (34, 36), headache (N = 2), tingling (N = 2), burning pain (N = 1), palpitation (N = 1) (41), and altered consciousness between treatment sessions (N = 1) (45).

## 4. Discussion

This systematic review demonstrates the promise of TBS in the treatment of AD, integrating evidence from both preclinical and clinical studies. Of particular importance is the potential of TBS to improve cognitive functioning, as most treatment modalities, at best, only slow the rate of decline. Preclinical studies demonstrated that TBS exerts neuroprotective effects by reducing Aβ burden, enhancing synaptic plasticity, alleviating neuroinflammation and oxidative stress, and improving cognitive function. In clinical studies, TBS, primarily in the form of iTBS or aiTBS targeting DLPFC, was associated with improvements in cognitive function, depression, anxiety, and activities of daily living. Functional and structural neuroimaging findings indicated that TBS modulates brain connectivity and preserves hippocampal volume. Notably, TBS was generally well-tolerated, with only minor adverse events, such as scalp discomfort and headaches reported.

Stimulation of different brain regions with TBS yielded distinct cognitive and neurophysiological outcomes. DLPFC-iTBS consistently improved global cognition, memory, and executive function while modulating network connectivity (33–35, 37, 38, 40, 44, 45). Specifically, iTBS improved functional connectivity between left DLPFC and bilateral caudate, as well as between the right amygdala and right caudate, regions implicated in spatial attention, working memory, and decision-making (48, 49). These effects may be mediated by excitatory modulation of the prefrontal cortex–a central hub in frontal-limbic circuits. TBS may also support neuronal function by enhancing cerebral metabolism, promoting cortical-subcortical interactions, and preserving structural integrity, contributing to cognitive improvements and symptomatic relief in patients with AD (35, 37). M1-iTBS, though less studied, highlighted impaired motor plasticity in AD, suggesting dysfunction in sensorimotor networks (39, 46). Moreover, LTP-like plasticity demonstrated moderate reproducibility among patients with AD, suggesting that the pathological processes underlying AD may simultaneously alter and stabilize TMS measures (46). However, variability in LTP-like plasticity among patients and protocols complicates interpretation of these findings. mPFC-iTBS enhanced cognition and mood, likely by modulating limbic-cognitive circuits, though its effects on executive function remains unclear (43). Additionally, TBS may impact motor and cognitive processes via cerebellar–cerebral pathways (50, 51), likely involving modulation of Purkinje cells and the dentate nucleus (52–56). However, findings from cerebellar TBS were mixed: cTBS suppressed, and iTBS enhanced, motor cortex excitability in healthy controls, whereas responses in patients with AD were blunted (41, 42). These findings highlight the region-specific neuroplasticity deficits in AD, with DLPFC-iTBS remaining the most promising target for cognitive enhancement. Building on this, comparisons among protocols further underscore how differences in stimulation parameters, such as intensity, frequency, spacing, and cortical target, significantly influence treatment efficacy. For instance, two studies administered high-frequency aiTBS over the left DLPFC with twice and three daily sessions, respectively, leading to notable improvements in global cognition and memory, with Wu’s regimen demonstrating sustained benefits at 10 weeks (34, 36). In contrast, Kashyap et al. (2024) employed a lower daily dose across individualized prefrontal targets and observed improvements limited to attentional domains (45). These findings underscore that TBS protocols vary in efficacy depending on stimulation parameters such as dosing schedule and target selection.

Optimizing these factors is crucial for achieving specific therapeutic outcomes, reinforcing the importance of individualized protocol design and precise anatomical targeting to maximize cognitive benefits in AD.

In preclinical studies, iTBS has been shown to mitigate oxidative stress, reduce Aβ and amyloid precursor protein levels, and decrease reactive astrogliosis (29). TBS exerts multifaceted effects by modulating neurotransmitter systems, particularly GABA and glutamate, and influencing the activity of NMDA and AMPA receptors (57–59). Furthermore, TBS enhances the expression of *BDNF*, a critical mediator of neuroplasticity and neuronal survival (60, 61). Notably, TBS has been associated with reduced expression of CD68 (27, 30), a marker of microglial activation involved in lysosomal Aβ degradation (62); thus, whether reduced CD68 expression should be interpreted as a favorable outcome of TBS requires further investigation. Collectively, these mechanisms underscore the therapeutic potential of iTBS in fostering neuroprotection and cognitive enhancement, positioning it as a potential strategy to decelerate AD progression (12).

Although preclinical studies robustly demonstrated the efficacy of TBS in animal models of AD, the findings from clinical studies were comparatively inconsistent and less conclusive. This discrepancy underscores the challenges in translating experimental success into clinical effectiveness and highlights the need for more rigorous human research. Moreover, although preclinical studies provided valuable mechanistic insights, the translational value of animal models is inherently limited. Rodent models only partially replicate the multifactorial pathology and chronic progression of human AD, and those induced by neurotoxins (e.g., STZ or TMT) may not accurately reflect sporadic AD pathophysiology.

Several limitations further constrain the interpretability and generalizability of the current evidence. First, the relatively small number of included studies and limited sample sizes, particularly in clinical trials, reduce statistical power. Several reported outcomes were derived from single studies with small cohorts and thus require replication in larger, well-powered, multi-center trials. Second, heterogeneity in study designs, including differences in study designs, TBS protocols (e.g., stimulation intensity, number of pulses, session frequency), targeted brain regions, and outcome measures, precluded quantitative meta-analysis and complicated cross-study comparisons. This variability also limits the identification of optimal stimulation parameters for specific therapeutic goals. Third, the predominant focus on the DLPFC may introduce bias, as the effects of stimulating other potentially relevant brain regions remain underexplored. Region-specific neuroplasticity responses, especially involving the cerebellum, motor cortex, and medial prefrontal cortex, were insufficiently studied, highlighting a gap in anatomical targeting research. Fourth, short follow-up periods in most studies also limit conclusions about the durability of therapeutic effects. Fifth, there was no consistent reporting on whether AD diagnoses were based on biomarker evidence or clinical judgment alone, a factor that significantly impacts the generalizability and interpretability of findings. Future large-scale, multi-center trials should focus on optimizing stimulation protocols, exploring individualized approaches, and assessing long-term clinical outcomes to advance the clinical application of TBS in AD.

## 5. Conclusions

TBS is a non-invasive neuromodulation technique that may hold potential for AD, with preliminary evidence suggesting possible benefits for cognitive function and neuroplasticity. Although the DLPFC is the most frequently targeted region, early findings from stimulation of other areas such as the mPFC, M1, and cerebellum warrant further exploration. Given that current conclusions are primarily based on a limited number of studies, in cases, single studies, more rigorous, large-scale research is needed to clarify the mechanisms, efficacy, and clinical relevance of TBS in AD.

## Supporting information

Supplementary material 1 (Search strategy)

## List of abbreviation

Aβ: Amyloid-beta
AD: Alzheimer’s disease
ADAS-Cog: Alzheimer’s Disease Assessment Scale-Cognitive Subscale
ADL: Activities of Daily Living
Akt: Protein kinase B
AMPA: α-amino-3-hydroxy-5-methyl-4-isoxazolepropionic acid
APP: Amyloid precursor protein
AVLT: Auditory Verbal Learning Test
BACE1: β-Site amyloid precursor protein cleaving enzyme 1
BDNF: Brain-derived neurotrophic factor
BNT: Boston Naming Test
CD68: Cluster of differentiation 68
CDR: Clinical Dementia Rating
CDT: Clock-Drawing Test
cTBS: Continuous theta burst stimulation
DLPFC: Dorsolateral prefrontal cortex
DST: Digit Span Test
ERK1/2: Extracellular signal-regulated kinases 1 and 2
GFAP: Glial fibrillary acidic protein
GMV: Gray matter volume
GSH: Glutathione
HAMA: Hamilton Anxiety Scale
HAMD/HDRS: Hamilton Depression Rating Scale
Iba1: Ionized calcium-binding adapter molecule 1
ICF: Intracortical facilitation
IDE: Insulin-degrading enzyme
IFN-γ: Interferon-gamma
IL-1β: Interleukin-1 beta
IL-6: Interleukin-6
iTBS: Intermittent theta burst stimulation
JBI: Joanna Briggs Institute
M1: Primary motor cortex
MDA: Malondialdehyde
MEP: Motor evoked potential
MnSOD: Manganese superoxide dismutase
MMSE: Mini-Mental State Examination
MoCA: Montreal Cognitive Assessment
mTOR: Mammalian target of rapamycin
NeuN: Neuronal nuclei
NMDA: N-methyl-D-aspartate
NPI: Neuropsychiatric Inventory
NO2−: Nitrite
PI3K: Phosphoinositide 3-kinase
PROSPERO: International Prospective Register of Systematic Reviews
PRISMA: Preferred Reporting Items for Systematic Reviews and Meta-Analyses
PSD95: Postsynaptic density protein 95
QoL-AD: Quality of Life in Alzheimer’s Disease
RBANS: Repeatable Battery for the Assessment of Neuropsychological Status
rMT: Resting motor threshold
rTMS: Repetitive transcranial magnetic stimulation
SCWT: Stroop Color-Word Test
SICI: Short-interval intracortical inhibition
SLAI: Short-latency afferent inhibition
SNAP25: Synaptosomal-associated protein 25
SYRCLE: Systematic Review Centre for Laboratory Animal Experimentation
SYN1: Synapsin-1
TBS: Theta burst stimulation
TMT: Trail Making Test
TNF-α: Tumor necrosis factor-alpha
tSOD: Total superoxide dismutase
VAMP1: Vesicle-associated membrane protein 1
VFT: Verbal Fluency Test.

## Declarations

### Ethics approval and consent to participate

Not applicable

### Consent for publication

Not applicable

### Availability of data and materials

Data sharing is not applicable to this article, as no datasets were generated or analyzed during the current study, which was conducted as a systematic review.

## Funding

The authors received no financial support for the research, authorship, and/or publication of this article.

### Authors’ contributions

Conceptualization: NE, HM and PA; Data curation: NE, HM and PA; Methodology: NE and HM; Project administration: NE, HM, and PA; Supervision: MSH and NR; Validation: MSH and NR; Writing - original draft; NE, HM and PA; and Writing - review & editing: NE, HM, PA, NR, and MSH. All authors have read and approved the submitted manuscript.

## Acknowledgements

The authors would like to thank Professor Mohammad Rohani for his invaluable support and insightful guidance.

## Supplementary Information

**Additional file 1**

**Title:** Search strategy and eligibility criteria for study selection

**Description:** This file provides the search strategy used to identify studies on Theta Burst Stimulation (TBS) for Alzheimer’s disease, including search terms, databases, and inclusion criteria.

