## Supplementary material 1 (Search strategy) for "Alzheimer’s disease and the therapeutic potential of theta burst stimulation: A systematic review of preclinical and clinical studies"

**PubMed/MEDLINE**

("Transcranial Magnetic Stimulation"[MeSH Terms] OR "TMS"[Title/Abstract] OR "Repetitive Transcranial Magnetic Stimulation"[Title/Abstract] OR "rTMS"[Title/Abstract] OR "Theta Burst Stimulation"[Title/Abstract] OR "iTBS"[Title/Abstract] OR "cTBS"[Title/Abstract] OR "Intermittent Theta Burst Stimulation"[Title/Abstract] OR "Continuous Theta Burst Stimulation"[Title/Abstract])

AND

("Dementia"[MeSH Terms] OR "dement*"[Title/Abstract] OR "Alzheimer Disease"[MeSH Terms] OR "Alzheimer*"[Title/Abstract])

**Scopus**

(TITLE-ABS("Transcranial Magnetic Stimulation" OR TMS OR "Repetitive Transcranial Magnetic Stimulation" OR rTMS OR "Theta Burst Stimulation" OR iTBS OR cTBS OR "Intermittent Theta Burst Stimulation" OR "Continuous Theta Burst Stimulation"))

AND

(TITLE-ABS(dement* OR "Alzheimer Disease" OR Alzheimer*))

**Cochrane library, trials**

("Transcranial Magnetic Stimulation" OR TMS OR "Repetitive Transcranial Magnetic Stimulation" OR rTMS OR "Theta Burst Stimulation" OR iTBS OR cTBS OR "Intermittent Theta Burst Stimulation" OR "Continuous Theta Burst Stimulation")

AND

("dementia" OR dement* OR "Alzheimer Disease" OR Alzheimer*)

**Embase**

('Transcranial magnetic stimulation'/exp OR 'transcranial magnetic stimulation':ti,ab OR tms:ti,ab OR 'Repetitive transcranial magnetic stimulation':ti,ab OR rtms:ti,ab OR 'Theta burst stimulation':ti,ab OR itbs:ti,ab OR ctbs:ti,ab OR 'Intermittent theta burst stimulation':ti,ab OR 'Continuous theta burst stimulation':ti,ab)

AND

('Dementia'/exp OR dementia:ti,ab OR dement*:ti,ab OR 'Alzheimer disease'/exp OR alzheimer*:ti,ab)

**WOS**

TS=("Transcranial Magnetic Stimulation" OR TMS OR "Repetitive Transcranial Magnetic Stimulation" OR rTMS OR "Theta Burst Stimulation" OR iTBS OR cTBS OR "Intermittent Theta Burst Stimulation" OR "Continuous Theta Burst Stimulation")

AND

TS=("Dementia" OR dementia* OR "Alzheimer Disease" OR Alzheimer*)
